# A New Advanced Osteoarthritis Treatment Utilizing Modified Mesenchymal Stem Cells: Arthroscopic Guided Intra-Articular Intervention Approach a Systematic Review and Meta-Analysis

**DOI:** 10.1101/2023.12.18.23299488

**Authors:** Kevin Christian Tjandra, Robin Novriansyah, Ardiyana Ar, Nurul Azizah Dian Rahmawati, I Nyoman Sebastian Sudiasa, Ismail Hadisoebroto Dilogo

## Abstract

**Background:** The mesenchymal stem cells (MSCs) is able to regenerate the cartilage defect caused by osteoarthritis (OA) to prevent permanent disability. Its efficacy may be even greater in combination with platelet-rich plasma (PRP) and hyaluronic acid (HA). Thus, this systematic review aimed to investigate the efficacy of MSCs in combination with PRP and adjusted doses of HA, the best source of MSCs, and the optimal number of applied MSCs to treat osteoarthritis as a cartilage regenerative agent.

**Method:** The sources included were original articles published from 2013 until 2023 from 4 databases (Pubmed, Springerlink, ScienceDirect, and Google Scholar). Studies included were original research of clinical trials or randomized controlled trials. Irrelevant studies were excluded. Then, the ROB-2 taken was used to assess bias. The result was constructed with PICOS criteria within the table created in the Google spreadsheet. MRI score, VAS score, Lysholm score, Cartilage volume, size of cartilage defect, Knee Society Clinical Rating System Score (KSS), and WOMAC index to evaluate treatment’s effication outcomes were analyzed by Revman 5.4. This systematic review followed the PRISMA guidelines.

**Result:** nine studies were included in the final screening. The meta-analysis showed a significant (P < 0.00001) elevation of Lysholm score with a pooled mean difference (MD) of (17.89) (95% CI: 16.01, 19.77; I^2^ = 0%, P = 0.56); a significant reduction (P < 0.00001) of VAS score with a pooled MD of (-2.62) (95% CI: -2.83, -2.41; I^2^ = 99%, P <0.00001); Knee society clinical rating system score (KSS) evaluation also showed significant elevation (P< 0.00001) with mean polled (29.59) (95% CI: 27.66, 31.52; I^2^= 95%, P< 0.0001); and significantly reduction (P<0.00001) of WOMAC score occurred as pooled MD of (-12.38) (95% CI: -13.75, -11.01; I^2^= 99%, P< 0.0001).

**Conclusions:** Arthroscopic guided high-dose subchondral application of primary cultured synovial mesenchymal stem cells in popliteal platelet-rich plasma media combined with hyaluronic acid effectively regenerate cartilage defect and increase clinical outcomes in the early stage of osteoarthritis.

**Level of Evidence:** Therapeutic Level I.

## Introduction

Osteoarthritis is a chronic inflammatory joint disease due to the degradation of the cartilage that causes joint pain and limitation in movement [1]. Over three decades, the global prevalence of OA increased by 113.25% from 1990-2019 with an estimated 527.81 million people [2]. The pathogenesis of OA is complex due to the multifactorial factors that can influence the occurrence and progression of the disease, including genetic, cellular, biochemical, and immunological. However, the main pathogenesis pathway that worsens the OA condition is the calcification of the cartilage defect. Thus, cartilage repair may prevent the progression of osteoarthritis [3][4].

The pharmacological conservative therapy does not provide significant benefits and the side effects that can arise also need to be considered in its application [5]. Furthermore, pharmacotherapy itself is not able to solve the main problem of OA, cartilage damage. Surgical techniques such as total knee replacement can be performed but the risk of persistent pain or loss of joint function reaches 20% [6]. Thus, less invasive procedure with promising result is needed to treat OA.

Mesenchymal stem cells (MSCs) as the regenerative agent is one of the solution of knee OA treatment because of its ability to regenerate cartilage defect [7]. Among various MSC sources, synovial MSC is a promising cell source for cartilage repair. Synovial MSCs have superior chondrogenic ability compared to MSCs from other tissues [8]. Synovial tissue has the greatest potential for differentiation into chondrogenic cells and proliferation. The synovium is a thin membrane that covers the inside of the joint and has a high regenerative activity. According to previous studies, the amount of MSCs in synovial fluid is increased in knees with OA that proving its regeneration activity [9]. Transplantation of large numbers of synovial MSCs into injured cartilage tissue can enhance the natural healing process. From previous studies, synovial MSCs were transplanted arthroscopically and led patients to return to daily life and sports activities earlier than patients with invasive surgery [8]. Therefore, this systematic review aimed to investigate the efficacy of MSCs in combination with PRP and adjusted doses of HA, the best source of MSCs, and the optimal number of applied MSCs to treat osteoarthritis as a cartilage regenerative agent.

## Material and Method

### Registration

The Preferred Reporting Items for Systematic Reviews and Meta-Analyses (PRISMA) were used for this systematic review [10]. On August 8, 2023, this systematic review and meta-analysis was registered to the Open Science Framework (OSF).

### Eligibility criteria

From 2013 until 2023 (the last date for which a search was conducted was 22 May 2023), we have included original content. The study included original research publications that met the inclusion criteria for autologous clinical trials and randomized controlled trials (RCT). Technical reports, editor answers, narrative reviews, systematic reviews, meta-analyses, non-comparative research, in silico studies, in vitro studies, in vivo studies, scientific posters, study protocols, and conference abstracts were all discarded. Non-English, non-full-text, and unrelated papers that were not related to the application of MSCs to treat knee osteoarthritis were also removed. The desired PICO criteria of the selected articles including i) patient with osteoarthritis or cartilage damage; ii) Intervention of intra-articular intervention or arthroscopic guided MSCs application; iii) Comparison, standard care without synovial stem cell application (placebo); and iv) Outcomes, treatment’s evaluation of AMADEUS MRI score for cartilage defect, VAS score, Lysholm score, Cartilage volume, size of cartilage defect, Knee Society Clinical Rating System Score (KSS), and WOMAC index.

### Outcome measure

The efficacy evaluation of the therapy was then assessed by several outcomes including MRI score, VAS score, Lysholm score, Cartilage volume, size of cartilage defect, Knee Society Clinical Rating System Score (KSS), and WOMAC index. The AMADEUS MRI score for cartilage defects was assessed before and after treatment to know the cartilage regeneration; the VAS score was used to assess the clinical pain before and after treatment; the Lysholm score was used to assess the symptom of disability; then the WOMAC index has a role to assess the severity clinical problem in osteoarthritis patients; and KSS is used to evaluate rate the knee and patient’s functional abilities.

### Index Test

Studies that provided data on the application and evaluations of bone membrane mesenchymal stem cells in osteoarthritis are included in this systematic review and meta-analysis.

### Reference Standard

Reference standards are professional research using randomized controlled clinical studies (RCT) or clinical trials to assess the transition of mesenchymal stem cells in osteoarthritis outcomes.

### Data Sources and Search

Research for this study was gathered through Pubmed, Springerlink, Google Scholar, and Science Direct database searches. The database was searched from its establishment until 22 May 2023 during 10-year periods prior to this review. The Boolean operator was utilized among the Medical Subject Headings (MeSH) keywords determined from the national institute of Health (NIH) National Library of medicine browser. In **Table 1**, the keywords used in each database are displayed. The studies are kept using Mendeley Group Reference Manager in the authors’ library. Clinical trial and randomized controlled trial type articles were used as a filter in the Pubmed database, the content-type article was used as the Springer database filter, the research article type was used for the Science Direct database filter, and any type of article was used in Google Scholar. A total of 5,101 studies were found in the database (4478 from Springerlink, 521 from Pubmed, 70 from Google Scholar, and 32 from Science Direct). 561 studies were imported to the Mendeley Group Reference Manager in the authors’ library after the aforementioned criteria were added, and this was done before the selection procedure.

**Table 1.**
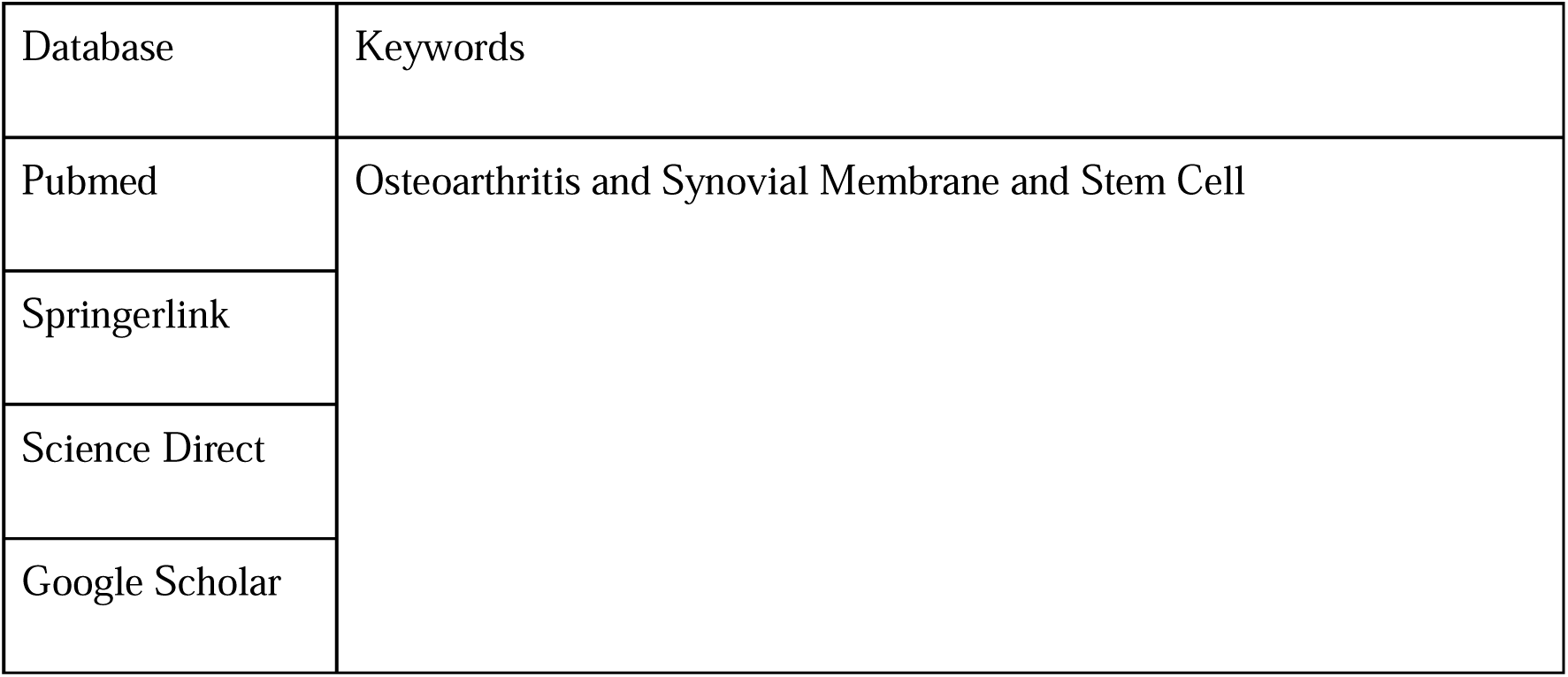
Keyword Used in Literature Searching.

### Selection process

Four independent reviewers (KCT, AA, NADR, and INSS) and one validator (RN) combined the outcomes from four databases after applying the search keywords indicated in Table 1. After that, they conducted a complete text and abstract screening to exclude non-clinical trial and non-randomized controlled trial papers while keeping the pertinent ones. 534 studies were eliminated from this process because they did not provide sufficient data to answer the research question (application and evaluations of MSCs in osteoarthritis) and because their study designs did not meet the criteria for inclusion (clinical trial and randomized controlled trial) and 16 studies were removed due to duplication. The retrieval of the complete text for the remaining 11 studies were then tested. Only 10 full text articles could be obtained as a result. Then, utilizing the title, year of publication, and DOIs, one publication was eliminated due to irrelevant outcome of interest. Using the Cochrane Risk of Bias 2 For Randomized Trials (ROB-2) tool, the nine included studies were evaluated for eligibility. All nine of the listed studies from this process passed the evaluation bias check. The PRISMA flow chart contained records of the research selection procedures.

### Data collection process

After the final screening, the pertinent information from studies is retrieved and entered into a Google Spreadsheet. Recorded datas in characteristic table consists of author, year, country, study design, sample size, gender, mean age, intervention name, comparison, and outcome consisting of the AMADEUS MRI score for cartilage defect, VAS score, WOMAC Index, Cartilage volume, size of cartilage defect, Knee Society Clinical Rating System Score (KSS).

### Study Risk of Bias assessment (Qualitative Synthesis)

Risk-of-bias tool used to assess the bias of included studies was the Cochrane risk-of-bias tool for randomized trials (RoB-2) which can be accessed in (https://methods.cochrane.org/bias/resources/rob-2-revised-cochrane-risk-bias-tool-randomized-trials), was used by five independent reviewers (KCT, RN, AA, NADR, INSS, and IHD). Any disagreements regarding the bias assessment were discussed further and settled between the 5 reviewers. Papers deemed with a high risk of bias will be removed from the systematic review to preserve the validity of included data in the current study as shown in.

### Reporting bias asseessment

As the result of ROB-2 assessment bias conducted by KCT, AA, NADR, INSS, and RN, all nine included articles did not show any critical overall bias as mentioned in Figure 1 so no article was removed at this stage.

**Figure 1.**
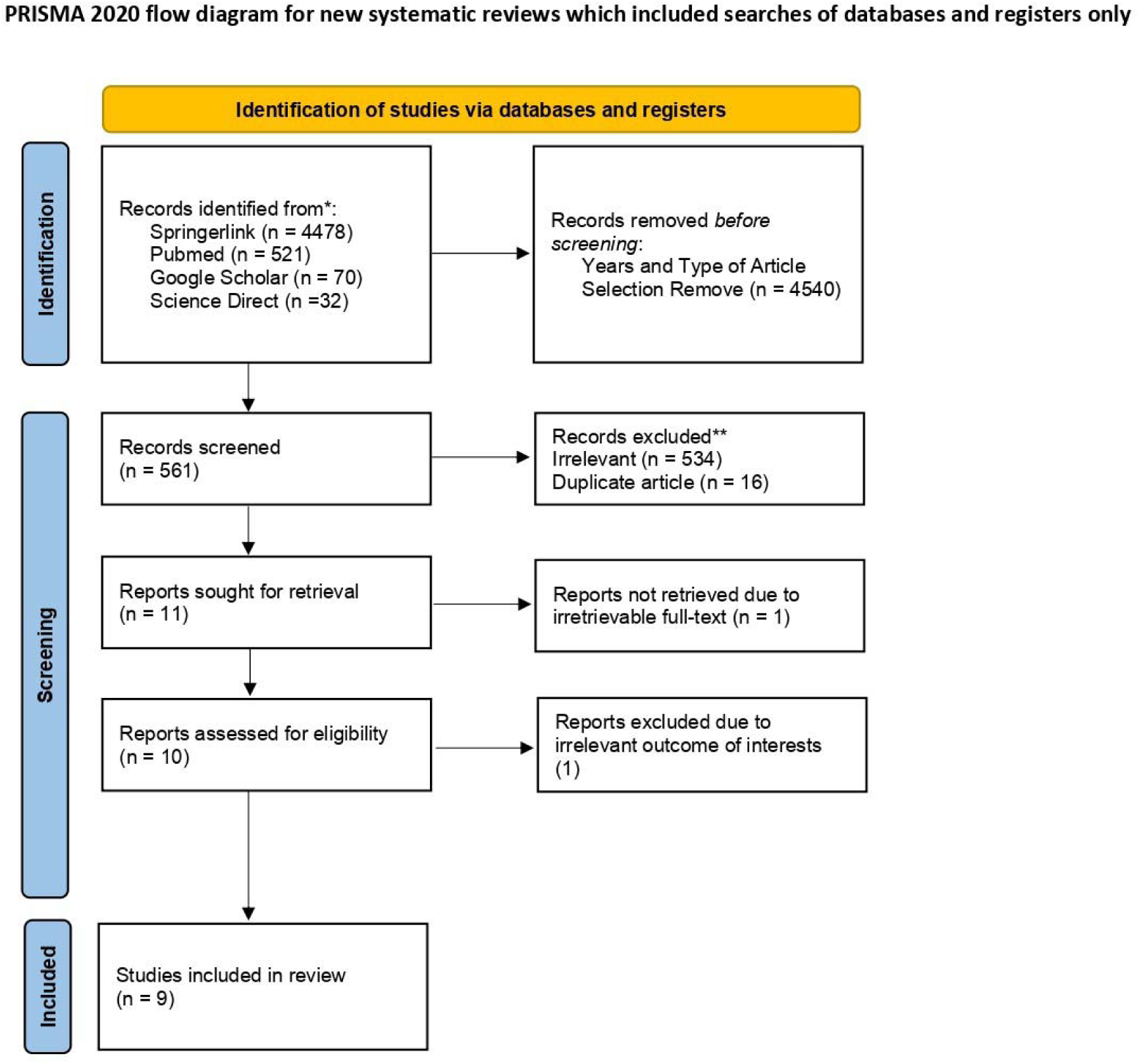
PRISMA 2020 flow diagram.

### Quantitative Synthesis

Outcomes were presented in the form of mean difference (MD) and standard deviation (SD) with a confidence interval (CI) of 95% in the characteristic table. Meta-analysis was used with a fixed-effect model (FEM) when the included studies were considered homogenous (low variability in studies’ results or variation due to random error), which was indicated by an I^2^ value of less than 40%. Otherwise, we used a random-effect model (REM). The pooled mean difference estimate was presented in a forest plot.

## Result

### Study Selection

A literature search utilizing Springerlink, Pubmed, Google Scholar, and ScienceDirect resulted in 5,101 papers overall. Automation approaches from each database were used to exclude non-clinical trials and non-randomized controlled trial research, which resulted in the elimination of 4,540 articles. After writers checked each and every article for relevancy and duplication starting with the title and abstract, 534 irrelevant subjects and 16 duplicate articles were eliminated. The inability to get the complete text document caused 1 unretrieved item to be eliminated. Then, 1 irrelevant outcome of interest research papers were eliminated. The full texts of nine articles were subsequently acquired. Last but not least, the author determined each study’s eligibility, and all nine studies met the requirements. This systematic review and meta-analysis include nine articles. The flow chart for the PRISMA diagram illustrating our research selection procedure is shown in Figure 1.

### Study Characteristics

The nine studies that made up this systematic review comprised a total of 315 participants. Each four studies were conducted in different countries. Studies were carried out in Japan, South Korea, France, China, USA, Iran, Singapore, and Spain. The complete study characteristics, including the PICO of each study, are stated in **Table 2**.

### Risk of Bias in Studies

Each clinical trial and randomized controlled trial study underwent a thorough assessment of its quality using the ROB-2 risk-of-bias method. In nine of the investigations, there were 3 studies identified as a study with some concern of bias due to unclear randomization process between the allocation of intervention and control group. An overview of the bias risk assessment is shown in Figure 2.

**Figure 2.**
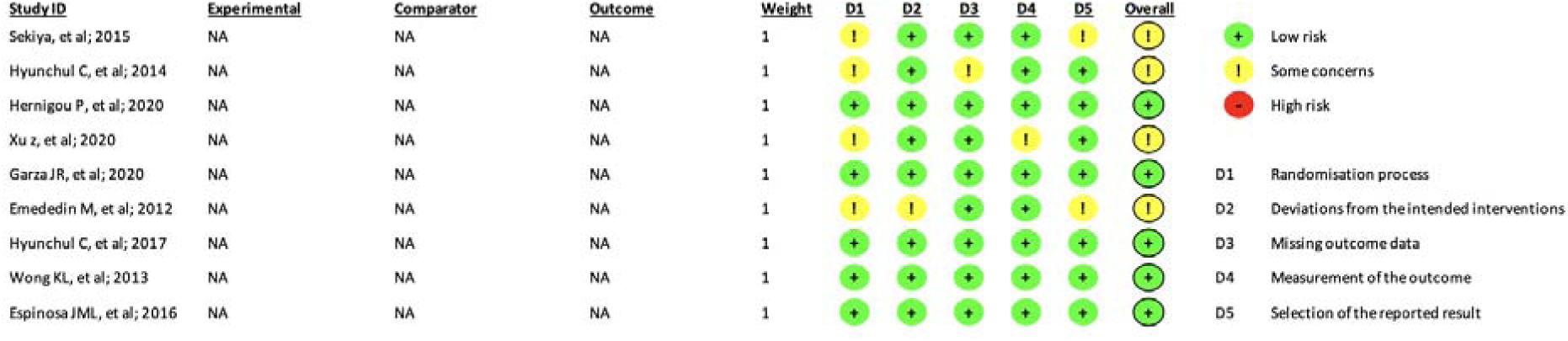
Risk of Bias Assessment

### Study Result Summaries

The journal selection method that has been carried out produced four studies that were used in the systematic review and meta-analysis process. The nine studies were conducted by Sekiya et al. (2015), Hyunchul C et al. (2014), Hernigou et al. (2020) Xu Z et al. (2020), Garza JR et al. (2020), Emededin M et al. (2012), Hyunchul C et al. (2017), Wong KL et al. (2013), Espinosa JML, et al. (2016). Brief profiles of the nine studies are shown in Table 2.

### Lysholm score

The disability symptoms caused by OA were assessed based on Lysholm’s score in the questionnaires of the respective standardized tools. A meta-analysis of two studies assessed the efficacy of OA synovial fluid stem cells in relieving disability symptoms in the pre-intervention and post-intervention groups. The forest plot in Figure 3 depicts a significant effect (P < 0.00001) with a pooled mean difference (MD) of (17.89) (95% CI: 16.01, 19.77). This result indicates that synovial fluid stem cell was found to decrease Lysholm score in OA patients significantly. The forest plot also showed heterogeneity was found (I2 = 0%; P <0.00001) for the efficacy of synovial fluid stem cells in relieving the disability symptoms in OA patients.

**Figure 3.**
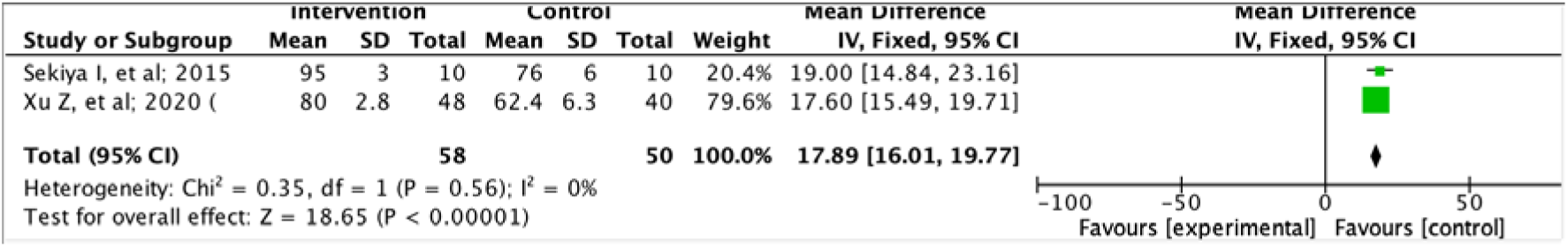
Forest plot of Lysholm score between intervention vs control group

### Visual Analog Scale (VAS)

Clinical pain experienced by OA patients is assessed with VAS scores which are shown in Figure 4. A meta-analysis included three studies that analyzed the efficacy of synovial fluid stem cells in reducing clinical pain in OA patients. The forest plot showed a significant effect (P < 0.00001) with a pooled MD of -2.62 (95% CI: -2.83, -2.41). The pooled MD shows that mesenchymal stem cell is significantly effective in reducing clinical pain in OA patients. Heterogeneity was found (I2 = 0%; P = <0.00001).

**Figure 4.**
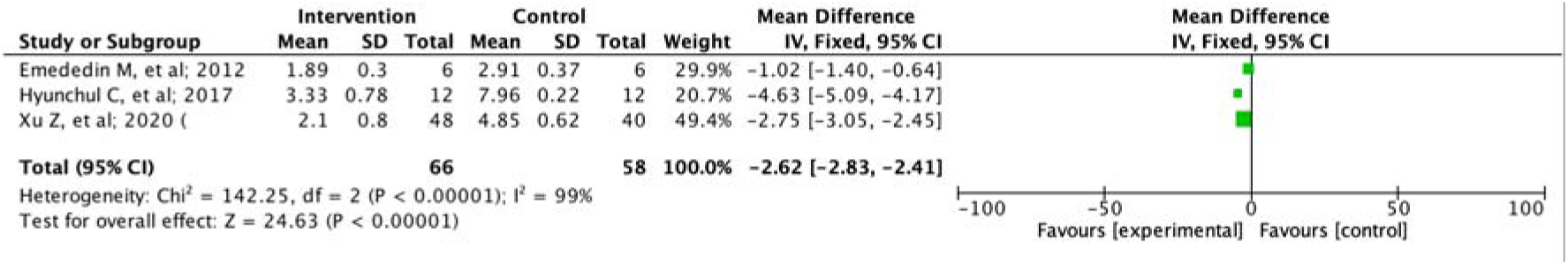
Forest plot of VAS score between intervention vs control group

### Knee Society Clinical Rating System Score (KSS)

Knee function and ability in OA patients is assessed with KSS scores as shown in Figure 5. The forest plot using KSS showed a significant effect (P < 0.00001) with a pooled MD of 29.59 (95% CI: 27.66, 31.52). The pooled MD shows mesenchymal stem cells significantly improved knee function and ability. Heterogeneity was found (I2 = 95%; P < 0.00001).

**Figure 5.**
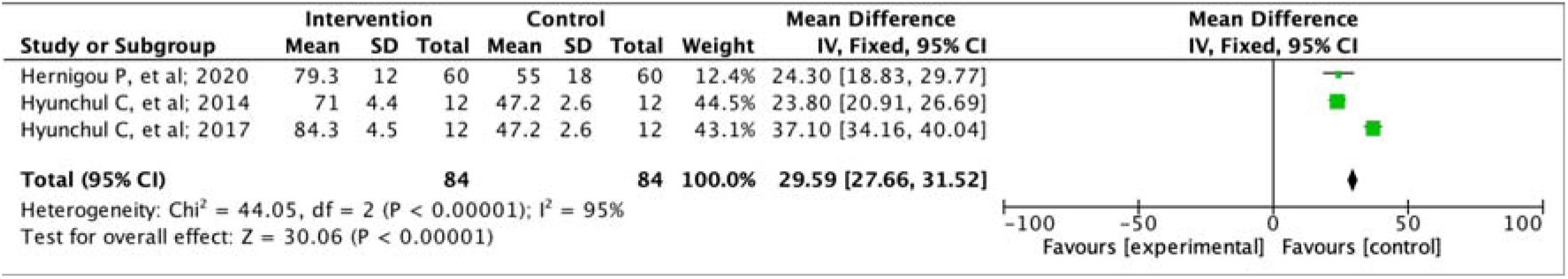
Forest plot of KSS score between intervention and control group.

### WOMAC Score

Symptoms in OA patients is assessed with WOMAC score as shown in Figure 6. The forest plot analyzing WOMAC score showed a significant effect (P < 0.00001) with a pooled MD of -12.38 (95% CI: -13.75, -11.01). The pooled MD shows mesenchymal stem cells significantly relieve symptoms in OA patients. Heterogeneity was found (I2 = 99%; P < 0.00001).

**Figure 6.**
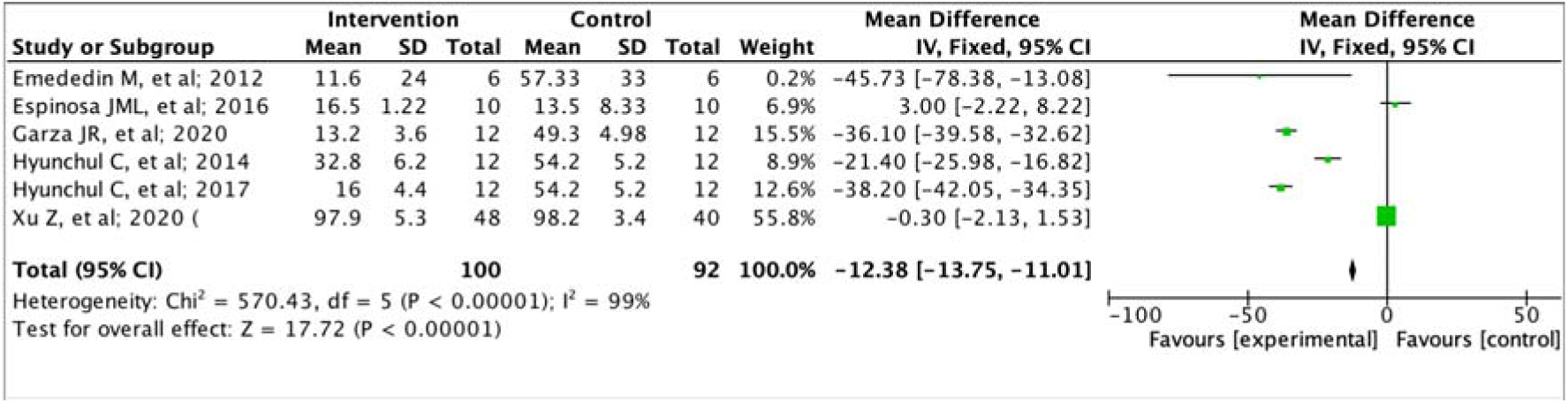
Forest plot of WOMAC score between intervention and control group.

## Discussion

### Synovial Mesenchymal Stem Cell

Synovial Mesenchymal Stem Cells (Sy-MSCs) originate from the mesenchyme, which is an embryonic connective tissue that gives rise to various types of cells, including bone, cartilage, and adipose tissue. Synovial MSCs have the ability to self-renew and differentiate into several types of cells from the mesenchymal lineage, including osteoblasts (bone cells), synovial (cartilage cells), and adipocytes (fat cells) [11]. They also exhibit immunomodulatory properties, meaning they can regulate the immune response. Synovial MSCs can be isolated from synovial fluid or the synovial membrane/tissue of joints such as the knee, hip, or shoulder. Various methods, including aspiration or biopsy, can be used to obtain these cells [12].

Synovial MSCs have garnered significant interest in regenerative medicine and tissue engineering due to their ability to differentiate into different cell types [13]. It has been shown that Sy-MSCs had a greater proliferation capacity and stronger chondrogenic potential than Bone Marrow-MSCs and Adipose Tissue-MSCs, as well as less hypertrophic differentiation than bone Bone Marrow-MSCs. In addition, pellet cultures were significantly larger from Sy-MSC than Bone Marrow-MSCs in patient-matched comparisons. They hold promise for the repair and regeneration of damaged or diseased joint tissues, such as cartilage and bone. They are being explored as potential treatments for conditions such as osteoarthritis, rheumatoid arthritis, and joint injuries [14].

Synovial MSCs play an important role in the pathophysiology and treatment of osteoarthritis (OA). In osteoarthritis, the synovial membrane undergoes changes, including inflammation and hyperplasia [15]. Synovial MSCs participate in the repair and maintenance of joint tissues, including articular cartilage. However, in OA, the regenerative capacity of synovial MSCs is often impaired due to factors such as aging, inflammation, and abnormal joint microenvironment. Synovial MSCs have chondrogenic potential, meaning they can differentiate into chondrocyte-like cells responsible for producing and maintaining the cartilage matrix [16][17][18].

Synovial MSCs hold promise for the treatment of OA. They can be isolated from synovial fluid or the synovial membrane through minimally invasive procedures [19]. These cells can be expanded in the laboratory and then transplanted back into the joint to promote tissue repair and regeneration.

### Application of Synovial Stem Cell in Treating Osteoarthritis

Osteoarthritis may develop as a result of articular cartilage injuries, which are a common clinical issue. Despite the variety of surgical intervention techniques, each has disadvantages of its own: Bone marrow stimulation results in poor structural quality of the repaired cartilage, donor site morbidity in mosaicplasty, and loss of the chondrogenic phenotype of expanded chondrocytes in autologous chondrocyte implantation [20]. One potential method for enhancing cartilage injury repair is stem cell therapy. Mesenchymal stem cells (MSCs), which can be isolated from various mesenchymal tissues, especially synovial membranes are one of the potential therapeutic cells.

The potential of synovial MSCs can be maximized by combining with other intraarticular regeneration agents, including, Platelet-Rich Plasma and hyaluronic acid [21]. This combination might be a great solution to treat cartilage damage in Osteoarthritis and prevent the overuse of hyaluronic acid that fastens the OA disease progression. A small amount of hyaluronic acid intraarticular application is able to stimulate cartilage regeneration. However, this small amount of HA is not enough to stop the disease progression, but the more hyaluronic acid is used the more VEGF production occurs. The high amount of VEGF will calcify the cartilage and convert it into osteocytes which worsen osteoarthritis [22].

Among all 4 Kellgern Lawrence classifications, this non-operative treatment is useful in OA grades 1 - 3. The earlier stage the greater outcome will be obtained [23]. It is because of this treatment strategy, that Modified Synovial Mesenchymal Stem Cells act as a cartilage regenerative agent that prevents further OA progression [8].

Cellaid1 (JMS Co Ltd, Hiroshima, Japan), a closed-bag system for serum isolation, was used to collect 300 mL of whole blood from the patient’s knee intraarticular, obtained by microfracture; fat pad severe; and suprapatellar pouch severe, one or two days prior to the harvest of synovial tissue. The popliteal vein is preferred due to its protein-rich plasma or autologous serum being suitable for the synovial stem cell culture process. The system consists of a blood donation bag filled with glass beads that, over 30 minutes of gentle mixing, activate platelets and remove fibrin from whole blood. The serum was isolated and heat-inactivated at 56° C for 30 minutes, following centrifugation at 2000 g for 7 minutes. Before use, the serum was stored at 4° C after filtering through a 0.45-lm nylon filter from Becton Dickinson, Franklin Lakes, New Jersey, USA [24].

The related Patients then underwent arthroscopy under the effects of local anesthesia containing 20 mL of 1% xylocaine. The synovium with sub synovial tissue on the femur at the suprapatellar pouch was then harvested with a pituitary rongeur under arthroscopic supervision while the patient was under intravenous anesthesia with 0.1 g sodium pentothal. After the synovial MSCs were harvested, patients were able to leave the hospital during the culture period [8].

The harvested synovial MSCs then digested using a solution of 5 mg Liberace in 5 ml Hanks’ balanced salt solution for 3 hours. Then the result was filtered through a 70-μm nylon filter to obtain the target synovial MSCs. The cells were grown in a-MEM from Invitrogen, which contained 10% autologous human serum, 100 units/mL of penicillin, 100 μg/mL of streptomycin, and 250 ng/mL of amphotericin B. In total, 50 dishes were examined on day 12 to look for bacteria, endotoxins in the medium, mycoplasma, and viruses in the cells. Chocolate agar was utilized for bacterial testing. A Toxicolor1 LS-50M kit (Seikagaku Corporation, Tokyo, Japan) was used for endotoxin testing. A multiplex PCR system created by our team was utilized for testing for mycoplasma and viruses [6]. This system detected 17 different virus types and 142 different mycoplasma types. Chromosomal testing was also conducted using one dish [25][26].

After no harmful contamination was confirmed, the synovial MSCs were harvested at day 14 using TrypLE at 37oC for 5 minutes. Primary synovial MSCs were suspended in 0.5 mL of acetate Ringer’s solution 30 minutes prior to transplantation. 47 million ± 21 million transplanted cells (mean ± SD) were used [27].

For the transplant, the patient was positioned with the cartilage defect facing upward, and using an 18-gauge needle and a 1-mL syringe, a suspension of synovial MSCs in 0.5 mL of acetate Ringer’s solution was injected into the defect under arthroscopic supervision. The defect was then held in the upward position for 10 minutes with the applied suspension. After 10 minutes, the additional 0.25 ml might be added to the cartilage defect in order to reduce synovial inflammation and reduce pain after the procedure [28].

Following surgery, all patients began knee ROM exercises. After two weeks and six weeks, they could bear total weight. There was no use of specialized machinery like a continuous passive motion machine. Generally, low-impact activities could begin at three months and high-impact activities at six. Then, the outcome evaluation was assessed using MRI, Lysholm score and Visual Analog Scale (VAS).

### Effication of MSCs in Treating Osteoarthritis

In recent years, the potential use of mesenchymal stem cells as a therapeutic approach for OA has gained significant attention. Compared to conventional bone marrow MSCs, Synovial mesenchymal stem cell (SFCs) has more efficacy as proven in the study conducted by Sekiya, et al. (2015) [8]. It’s because synovial fluid cells have more similarity in synovial environment than other sources of cells. Thus, it can exhibit multipotency where each cell can differentiate into various cell types including chondrocytes, which are crucial for the maintenance and repair of cartilage. The regenerative properties of SFSCs are important in a joint inflammation disease such as osteoarthritis. A study by Kim et al. (2019) demonstrated that SFSCs derived from OA patients could differentiate into chondrogenic lineage cells and promote cartilage matrix formation [29]. Moreover, another study by Qiu et al. (2021) reported that SFSCs combined with hyaluronic acid injections resulted in improved cartilage regeneration and enhanced functional outcomes in a rabbit model of OA [30]. Then, the efficacy of this treatment can be maximized by the application of appropriate high-dose MSCs (1 x 10^8^ cells) with subchondral technique compared to intra-articular injection as proven by a study conducted by Hernigou P, et al. (2020)[31].

However, studies exploring the efficacy of SFSCs in human trials are still relatively limited. A phase I/II clinical trial conducted by Jo et al. (2019) involved intra-articular injection of SFSCs in patients with knee OA [32].

This study reported significant improvements in pain, function, and cartilage quality, demonstrating the potential of MSCs as a new treatment modality for OA. Studies conducted by Sekiya et al. (2015) and Xu, et al. (2020) has proven elevation of the Lysholm score va which assesses disability in OA patients proved to be statistically significant (P <0.001) increased by polled MD (17.89) [95% CI:16.01, 19.77] compared to the control group [8] [33].

In addition, MSCs were also proven to be significantly effective in reducing VAS scores as proven by studies conducted by Emededin M, et al. (2012), Hyunchul, et al. (2017), and Xu Z, et al. (2020) showed the statistical effectiveness of Intra-articular Injection of Autologous MSCs (P <0.00001) with pooled MD (-2.62) (95% CI: -2.83, -2.41) [34] [35] [33]. Proven by MRI images, post-injection cartilage thickness increased in three of the six patients. Xu et al. (2020) showed that the efficacy of intra-articular platelet-rich plasma (PRP) SFSC would increase when combined with hyaluronic acid and (PRP) as demonstrated by a significant reduction in VAS scores in the PRP+HA group at 24 months afterward. injection, PRP+HA was more effective than HA and PRP alone in relieving pain (P = 0.0001) [33]. In addition, significant improvements were also observed in the PRP+HA group after 1, 6, 12, and 24 months (P < 0.0001).

MSCs can also improve the WOMAC index which assesses the severity of clinical problems in osteoarthritis patients. The results of the study by Emededin, et al. (2012), Espinosa, et al. (2016), Garza, et al. (2020), Hyunchul, et al. (2014), Hyunchul, et al. (2017), and Xu, et al. (2020) showed a clinically meaningful reduction of WOMAC score as much pooled MD (- 12.38) (95% CI: -13.75, -11.01) [34][36][37][38][35]. In addition, KSS function score evaluation from Hernigou, et al. (2020), Hyunchul, et al. (2014), and Hyunchul, et al. (2017) increased significantly from as much pooled MD (29.59) (95% CI: 27.66, 31.52) [31][35][38].

### Strength and Limitation

This study is the first systematic review and meta-analysis that assessed the efficacy of synovial fluid stem cell in treating osteoarthritis. This study also included studies from various countries showing this therapy’s universal applicability. Although this study also has several limitations such as the limited number of included studies.

## Conclusion

Arthroscopic guided injection of primary cultured synovial mesenchymal stem cells in popliteal platelet-rich plasma media combined with hyaluronic acid is effectively regenerate cartilage defect in the early stage of osteoarthritis and reduce synovial inflammation that may cause knee pain. Synovial mesenchymal stem cells have also been proven to not cause any side effects in osteoarthritis patients.

## Recommendation

More studies need to be conducted to further evaluate the duration and efficacy of the synovial mesenchymal stem cell as a novel therapy for osteoarthritis.

## Data availability

### Underlying data

All data underlying the results are available as part of the article and no additional source data are required.

### Reporting guidelines

Mendeley Data: A New Advanced Osteoarthritis Treatment Utilizing Modified Synovial Mesenchymal Stem Cells: Arthroscopic Guided Intra-Articular Intervention Approach a Systematic Review and Meta-Analysis. doi: 10.17632/zn8t3gks5g.

Data are available under the terms of the <u>Creative Commons Attribution 4.0 International license</u> (CC-BY 4.0).

