## Supplementary material for "A New Advanced Osteoarthritis Treatment Utilizing Modified Mesenchymal Stem Cells: Arthroscopic Guided Intra-Articular Intervention Approach a Systematic Review and Meta-Analysis": Table 1

**Table 1.** Keyword Used in Literature Searching

| Database | Keywords |
| --- | --- |
| Pubmed | Osteoarthritis and Synovial Membrane and Stem Cell |
| Springerlink |  |
| Science Direct |  |
| Google Scholar |  |
